# Non-Markovian modelling highlights the importance of age structure on Covid-19 epidemiological dynamics

**DOI:** 10.1101/2021.09.30.21264339

**Authors:** Bastien Reyné, Quentin Richard, Camille Noûs, Christian Selinger, Mircea T. Sofonea, Ramsès Djidjou-Demasse, Samuel Alizon

**Affiliations:** MIVEGEC, Univ. Montpellier, CNRS, IRD — Montpellier, France; Laboratoire Cogitamus; Swiss Tropical and Public Health Institute (Swiss TPH) — Basel, Switzerland; Center for Interdisciplinary Research in Biology (CIRB), College de France, CNRS, INSERM, Université PSL — Paris, France

**Keywords:** epidemiology, infectious diseases modelling, contact matrix, partial differential equation, Covid-19

## Abstract

The Covid-19 pandemic outbreak was followed by a huge amount of modelling studies in order to rapidly gain insights to implement the best public health policies. Most of these compartmental models involved ordinary differential equations (ODEs) systems. Such a formalism implicitly assumes that the time spent in each compartment does not depend on the time already spent in it, which is at odds with the clinical data. To overcome this “memoryless” issue, a widely used solution is to increase and chain the number of compartments of a unique reality (*e*.*g*. have infected individual move between several compartments). This allows for greater heterogeneity and thus be closer to the observed situation, but also tends to make the whole model more difficult to apprehend and parameterize. We develop a non-Markovian alternative formalism based on partial differential equations (PDEs) instead of ODEs, which, by construction, provides a memory structure for each compartment thereby allowing us to limit the number of compartments. We apply our model to the French 2021 SARS-CoV-2 epidemic and, while accounting for vaccine-induced and natural immunity, we analyse and determine the major components that contributed to the Covid-19 hospital admissions. The results indicate that the observed vaccination rate alone is not enough to control the epidemic, and a global sensitivity analysis highlights a huge uncertainty attributable to the age-structured contact matrix. Our study shows the flexibility and robustness of PDE formalism to capture national COVID-19 dynamics and opens perspectives to study medium or long-term scenarios involving immune waning or virus evolution.

## Introduction

Shortly after the Covid-19 outbreak in late 2019, many efforts were put in diverse research areas to understand both the disease and its aetiological agent, SARS-CoV-2, and to produce tools to deal with what quickly became a pandemic. Among those areas, mathematical modelling studies proliferated to better grasp the epidemics’ dynamics on a —at first— short and medium-term scale. Stochastic models were more appropriate early on to take into account the randomness of the transmission events (Beneteau et al., 2021; Hellewell et al., 2020; Kucharski et al., 2020), but they were rapidly complemented by deterministic models since many epidemics were settled within a couple of months in many countries. These modelling results provided valuable insights to guide public health policies, often on a nationwide scale (RSTB, 2021).

Many of the deterministic models developed were variations of the classical Susceptible– Infected–Recovered (SIR) compartmental model and usually involved a system of ordinary differential equations (ODEs) (Keeling and Rohani, 2008). Such a simple formalism was adapted at first given the unknowns regarding the natural history of the disease. However, the knowledge accumulated in a matter of months made it clear that several assumptions were biologically unrealistic. In particular, the residence times in various compartments were not distributed exponentially, and the related “lack of memory” (also named Markov property) —meaning that the time spent in a compartment does not depend on the time already spent in the compartment, as implicitly assumed by the ODE formalism— became particularly detrimental to short-term forecasting (see Supplementary Figure S1 for an illustration of the impact of memory on the time spent in a compartment). For example, for the time elapsed between infection and potential hospital admission, French hospitalisation data analyses show this memoryless hypothesis does not hold (Salje et al., 2020; Sofonea et al., 2021). A simple workaround to this issue consisted in adding new compartments, *e*.*g*. for exposed people but not yet infectious, thereby increasing the heterogeneity in the infected period. Earlier studies indeed show that the addition of intermediate compartments transforms the sum of exponentially distributed waiting times into a hypoexponential distribution (Lloyd, 2001). Accounting for memory can also be achieved by other formalisms such as discrete-time modelling, and thus be tailored to epidemiological data the time resolution of which is almost always that of the day (Sofonea et al., 2021).

Nevertheless, depending on the issue dealt with by the model, sources of heterogeneity may emerge and increase the number of host categories and thus the total number of equations and parameters. With the onset of vaccination campaigns, this phenomenon became even more pronounced (Kiem et al., 2021; Moore et al., 2021). Even if the approach consisting in a chain of compartments in ODE systems remained a useful approximation, the initial gain in simplicity progressively vanished, making the models increasingly difficult to analyse and parameterize.

On a longer time scale, virus evolution and the emergence of variants of concern (VOC) (Davies, Abbott, et al., 2021), coupled with some pre-existing unknowns regarding the behaviour of natural and vaccine-induced immune responses (Alizon and Sofonea, 2021; Zellweger et al., 2020), further increased the need for modelling approaches. However, even for medium or long-term projections, ODE-based approaches remain far from ideal since immunity waning may occur rapidly (at least from a prospective point of view) and might not be memoryless.

To address these issues, we use an alternate formalism relying on partial differential equations (PDEs) instead of ODEs, with which it shares similarities and simplicity in its formalism. Although models based on PDEs require an additional initial effort for parameterization, they offer increased flexibility because biological assumptions can be strongly varied without adding more compartments. Interestingly, the seminal work on the SIR model by Kermack and McKendrick (1927) was implicitly based on a PDE formalism and ODE models were simply presented as special cases when infectivity and removal rates were assumed to be constant. PDEs are often used in epidemiological models to take into account a population age-structure or a spatial structure (Brauer et al., 2019; Hethcote, 2000). PDE models can also elegantly incorporate non-linearity in models (*e*.*g*. for infectiousness profile (Hoppenstaedt, 1975; Inaba, 2017) or immunity waning (Ehrhardt et al., 2019)). Including such aspects with ODE-based models would require more effort because it would be necessary to add compartments and, therefore, change the structure of the model itself. Regarding Covid-19 epidemics, PDE based approached have mainly been used to deal with spatial structure (*e*.*g*. (Viguerie et al., 2021; Wang et al., 2020)) and sometimes to add the age of infection in the model structure (Richard et al., 2021; Wu et al., 2022). But, overall, PDE-based models of Covid-19 epidemics remain marginal compared to ODE-based models.

In this study, we build upon earlier work by Richard et al. (2021), who developed a non-Markovian model structured in terms of the age of the host (in years) and of the infection (in days). We extend it by generalising the PDEs non-linear properties to vaccination status and clearance memory. More precisely, we also record the time since vaccination as well as the time since clearance (both in days). These daily time structures allow us to keep track of the time spent in each compartment, thereby providing a convenient way to correct the memory problem while limiting the number of epidemiological compartments.

We first present the details of the model and derive some of its main properties, such as the basic reproduction number. Then, we perform a sensitivity analysis to identify which parameters affect epidemiological dynamics the most. We also show that the model can be tailored to analyse the past French Covid-19 epidemic and formulate projections regarding future trends. Finally, we discuss perspectives to extend the model and additional questions regarding Covid-19 epidemics.

## Model

### Model overview

The density of susceptible individuals of age *a* ∈ [0, *a*_max_] at time *t* is denoted by *S*(*t, a*). Susceptible individuals leave the compartment either by being infected, at a rate *λ*(*t, a*) corresponding to the force of infection, or by becoming vaccinated, at a rate *ρ*(*t, a*).

Infected individuals are denoted *I*^*ℓ*^(*t, a, i*). The exponent *ℓ* ∈ {*m, s, d*} corresponds to the three types of infections: *m* for mild and asymptomatic cases, *s* for severe cases that will require hospitalization at some point before recovering, and *d* for severe cases that always result in the patient’s death. Each of these categories of infected individuals is further stratified according to the time since infection, which is indexed by *i* ∈ [0, *i*_max_]. Practically, this affects the recovery rates (*γ*^*m*^(*a, i*) and *γ*^*s*^(*a, i*)) and the death rate (*μ*(*a, i*)), as well as the different transmission rates *β*^*ℓ*^(*a, i*), *ℓ* ∈ {*m, s, d*}; all of which are functions of *i*. The number of new mildly infected individuals at a given time *t* is given by the boundary condition,

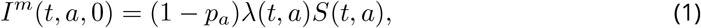

where *p*_*a*_ is the proportion of infections that lead to severe cases for individuals of age *a*.

We add a similar time structure *j* to record time since clearance for the density of recovered individuals, *R*(*t, a, j*), to account for a possible post-infection immunity waning at a rate *σ*(*a, j*). Recovered individuals are assumed to be vaccinated at the same rate as susceptible individuals, *ρ*(*t, a*). The number of newly recovered individuals of age *a* at time *t* is given by the boundary condition

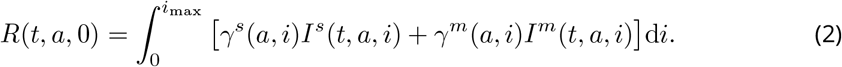

The density of vaccinated individuals, *V* (*t, a, k*), also has its own time-structure *k* to capture the time since vaccination. This allows taking into account the immunity waning, *σ*^*v*^(*a, k*), or any temporal variation in vaccine efficacy. The number of newly vaccinated individuals is given by the boundary condition

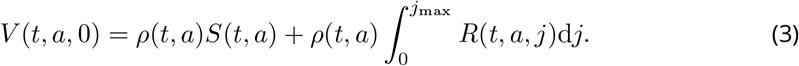

Since vaccine efficacy may be imperfect, we assume that vaccinated individuals can still be infected by the virus, but at a rate reduced by 1 − *ε*(*a, k*) compared to susceptible unvaccinated individuals. If the infection is mild, infected vaccinated hosts move to the *I*^*mv*^(*t, a, i, k*) compartment, which is separated from mild-infected former susceptible individuals to allow for reduced transmission at a rate 1 − *ξ*(*a, k*). Vaccinated individuals can also develop a severe form of Covid-19 following infection but at a rate reduced by (1 −*ν*(*a, k*)). And therefore reduced by (1−*ε*(*a, k*))(1−*ν*(*a, k*)) compared to susceptible individuals. Hence, the number of newly severely infected individuals of age *a* at time *t* is given by the boundary conditions

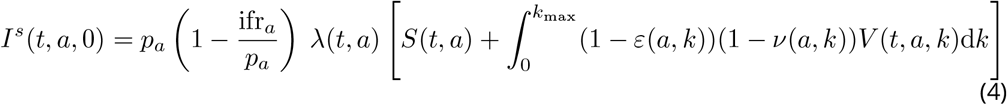

and

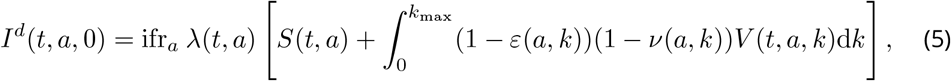

where ifr_*a*_ denotes the infection fatality rate (IFR), that is the fraction of individuals of age *a* who die from the infection. It is worth noting that due to VOC emergence inducing an increase in virulence, both *p*_*a*_ and ifr_*a*_ will be scaled by *κ* accounting for this increase.

Regarding the infected vaccinated individuals who develop mild symptoms, the boundary conditions are

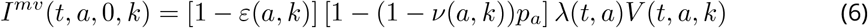

and

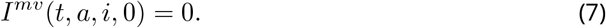

The model flowchart is displayed in Figure 1. Notice that our model only has 8 compartments. Note also that vaccines can act in three non mutually exclusive ways by decreasing the risk of being infected (*ε*(*a, k*)), the probability to develop severe symptoms if infected (*ν*(*a, k*)), and the transmission rate if infected (*ξ*(*a, k*)).

**Figure 1.**
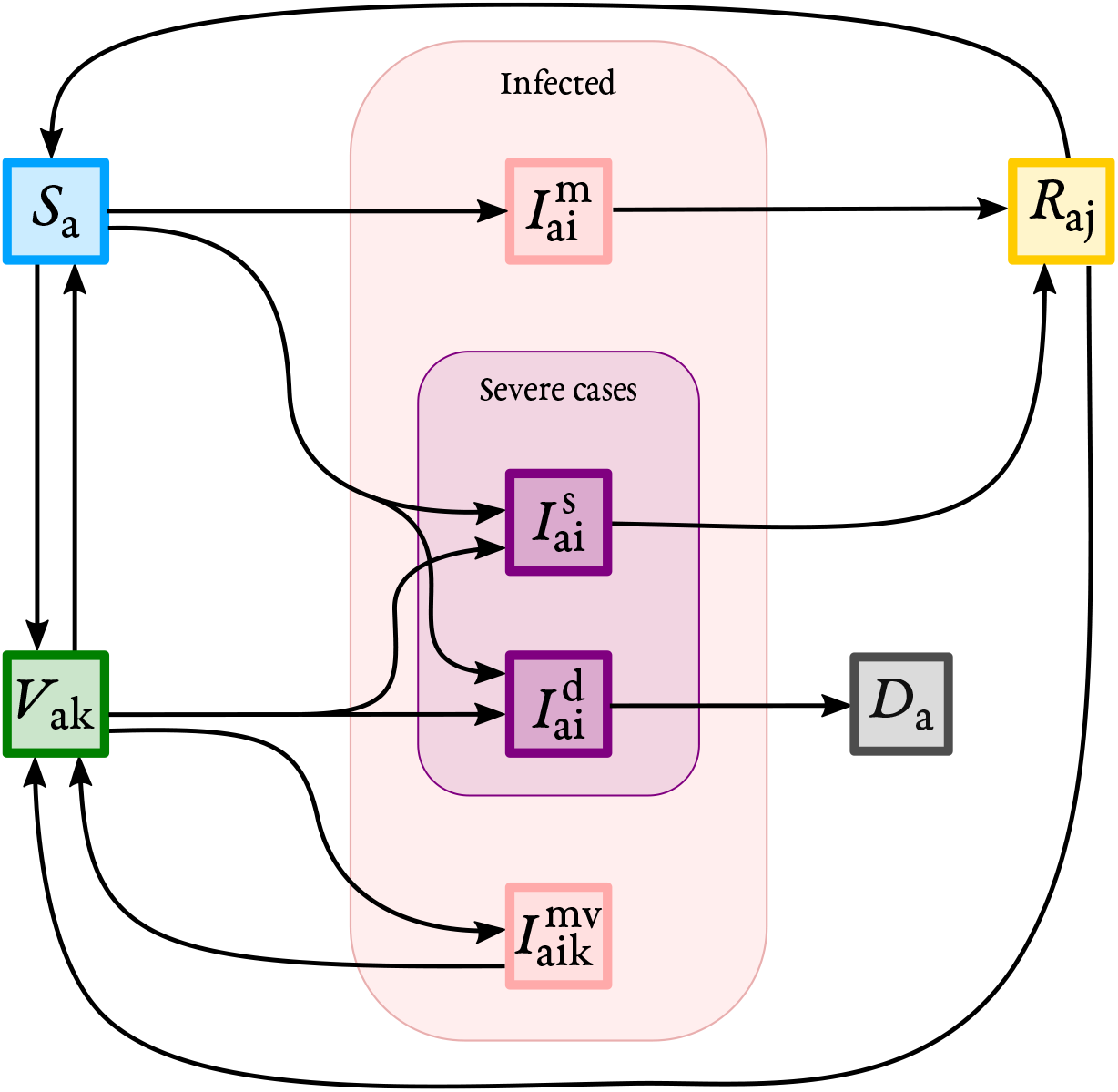
Model flowchart. On this flowchart, subscripts denote additional structure beside time *t* for each compartment: *a* stands for the host age, *i* for the time since infection, *j* for the time since clearance, and *k* for the time since vaccination. Exponents indicate different types of infections: *m* for mild or asymptomatic cases, *s* for severe cases who will survive, *d* for cases who will die, and *mv* for mild infection though vaccinated host.

The change over time, including the leaving of each compartment, is provided by the following PDE system, coupled with the boundary conditions (1)–(7):

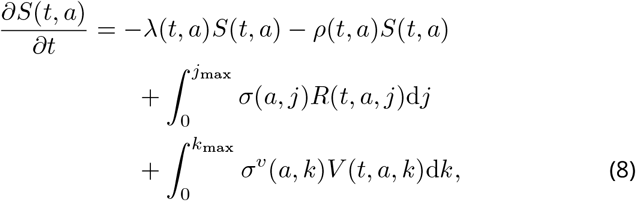

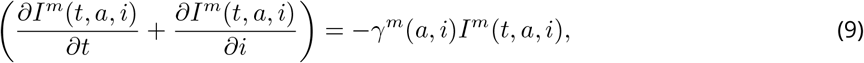

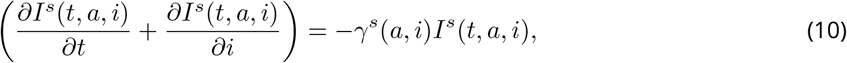

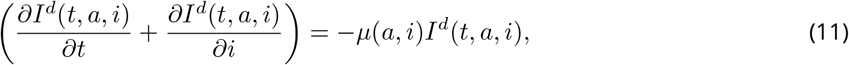

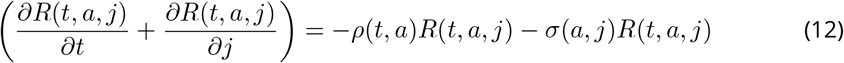

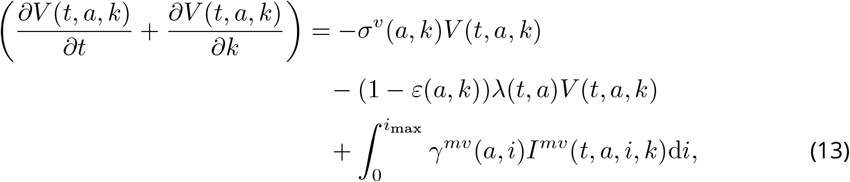

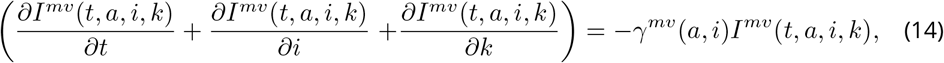

with

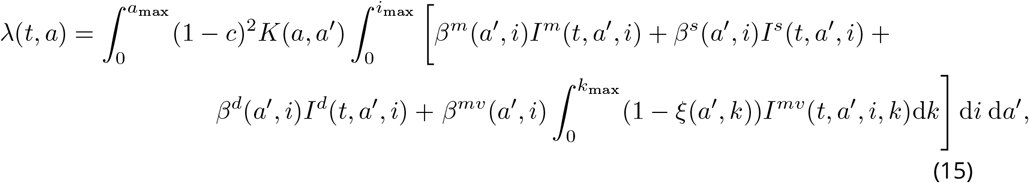

for any (*t, a, i, j, k*) ∈ ℝ^+^ × [0, *a*_max_] × [0, *i*_max_] × [0, *j*_max_] × [0, *k*_max_]. Here, *K*(*a, a*^′^) is the kernel giving the mean contact rate between two individuals belonging respectively to the age classes *a* and *a*^′^. We also introduce efficacy, denoted *c*, of non-pharmaceutical interventions (NPIs) in reducing the contact rates between individuals independently of their age. We assume NPIs affect all individuals indifferently, no matter the compartment they belong to. Therefore, since both susceptibles and infected individuals are targeted, the reduction of the contact rate is a squared term.

The above system is associated with Assumption S1 in Supplementary Methods and the following initial conditions

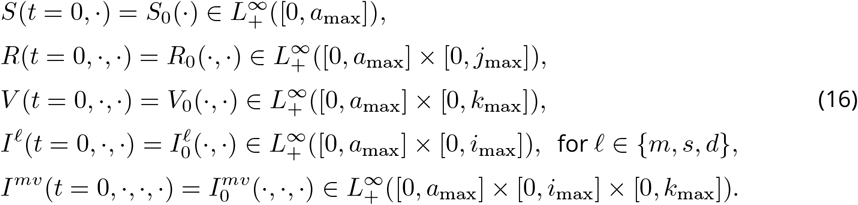

Notice that the well-posedness of System (1)–(16) is analysed in Supplementary Methods A.2.

### Model parametrization

In this study, we focus on the French Covid-19 epidemic in 2021. The values used are shown in Table 1 along with the (French) data we use for parameter inference. Additional details about these can be found in Supplementary Methods B.

**Table 1.** Model parameters. For each parameter, we indicate the default value used and the references used.

| Parameter | Value | Reference |
| --- | --- | --- |
| Generation time | Weibull(2.83, 5.67) | Ferretti et al. (2020) |
| Proportion of severe cases ($p_a$), IFR ($\text{ifr}_a$) and increase in virulence ($\kappa$) | 0.0113 (mean), 0.0022 (mean) and 1.65 (baseline) | Challen et al. (2021), Davies, Jarvis, et al. (2021), and Verity et al. (2020) |
| Mild recovery rate | Adapted | Salje et al. (2020) |
| Severe recovery rate | Adapted | Lefrancq et al. (2021) and Salje et al. (2020) |
| Vaccination rate | Fitted | https://data.gouv.fr |
| Total severity reduction | 0.925 (efficacy after 2 doses) | Public Health England (2021) |
| Infection immunity | 0.875 (efficacy after 2 doses) | Public Health England (2021) |
| Transmission reduction | 0.75 (efficacy after 2 doses) | Public Health England (2021) (see Supplementary Methods B.5) |
| Initial proportion of recovered | 0.149 [0.132 – 0.169] | Hozé et al. (2021) |
| Age structure | Real data | https://www.insee.fr/fr/statistiques/2381474 |
| Contact matrix | — | SPF/CNAM and Béraud et al. (2015) |

The basic reproduction number is fixed but varies in time due to the emergence of the *α* and *δ* VOCs. The *α* VOC was first detected in France in early January and rapidly became dominant. Therefore, the ℛ_0_ retained starting in January was 4.5 (Davies, Abbott, et al., 2021; Haim-Boukobza et al., 2021). By July, the *α* VOC was supplanted by the *δ* VOC, increasing the R_0_ up to 6 (Alizon, Haim-Boukobza, et al., 2021).

Regarding the modelling of vaccine efficacy, for simplicity, we neglect immune waning, *i*.*e*. the decrease of immunity with time, meaning that *σ*(*a, j*) ≡ 0 and *σ*^*v*^(*a, k*) ≡ 0. This assumption is motivated by the fact that we consider a medium-term scenario and it could readily be modified. We also assume that the three types of vaccine efficacies (against reinfection, severe symptoms, and transmission) are not maximal upon entry into the vaccinated compartment. More precisely, we assume a double sigmoid curve to capture two vaccine injections (Figure S2). The different levels of efficacy are based on the Public Health England (2021) report, and additional details are provided in the Supplementary Methods B.5. The vaccination rate *ρ*(*t, a*) is based on the observed French data (see Supplementary Methods B.6 for details about this implementation).

The different transmission rates *β*^*ℓ*^(*a, i*), *ℓ* ∈ {*m, s, d*}, are simply the generation time, weighted to correct for the possibility for individuals to leave the infected compartments before the generation time becomes null.

Concerning some age-stratified parametrization functions, we assume no differences between age groups. This assumption is either made for parsimony reasons (*i*.*e*. for *γ*^*m*^(*a, i*), *γ*^*s*^(*a, i*), and *μ*(*a, i*)) or because of lack of information (*e*.*g*. for *β*^*m*^(*a, i*) and *β*^*s*^(*a, i*)).

### Contact matrices

Due to the data available and following the parametrization relative to the severity disease, the kernel *K*(*a, a*^′^) is also given for a finite number of age classes, thus providing a contact matrix. And this contact matrix *K*(*a, a*^′^) is also an important part that needs to be defined as it will define the age-structure of the population regarding an age-severity differentiated infectious disease (Valle et al., 2013; Wallinga, Teunis, et al., 2006). In that regard, we decide to present two competing choices. The first one from Béraud et al. (2015) was estimated to better apprehend the spread of infectious diseases. The second source of contact matrices comes from the French health agency (Santé Publique France) and the French national health insurance (CNAM). They provide Covid-19 38 week-specific contact matrices ranging from August 2020 to April 2021.

The latter reveal pronounced changes across weeks. These are most likely due to a variety of reasons such as control restrictions policies or school holidays. Interestingly, these changes do not affect all age classes in the same way (Figure 2).

**Figure 2.**
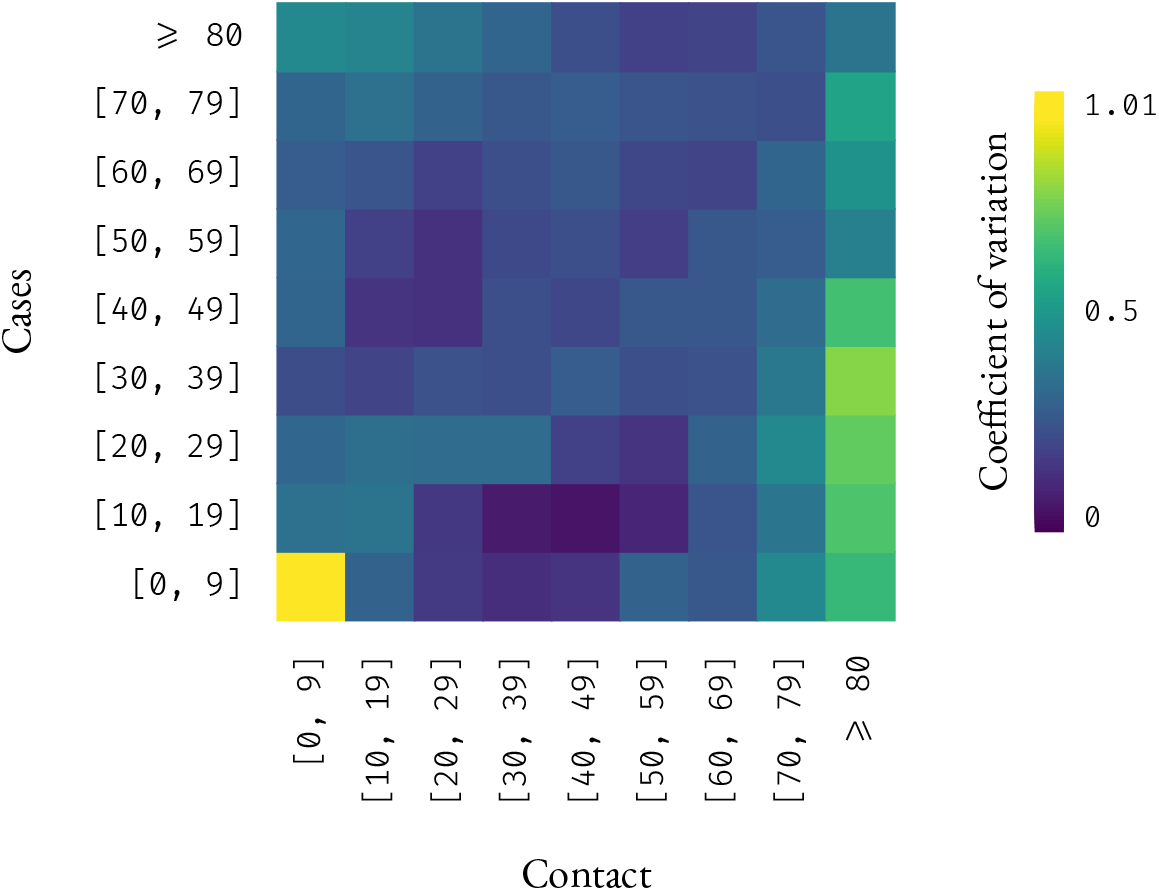
Coefficients of variation of each element of the SPF/CNAM contact matrix over the 38 weeks available. The higher the value, the greater the variability in contacts between age-classes over the different weeks.

The two sources of contact matrix also exhibit qualitative pattern differences, as illustrated in Figure 3. Indeed, that from Béraud et al. (2015) gives more weight to relatively young people who tend to have contact with people close in age, such as colleagues or friends, which could be representing the active population. Furthermore, in this matrix, older people have few contacts. Conversely, SPF matrices seem to have more extra-generational contacts, which could increase the role of transmission within households.

**Figure 3.**
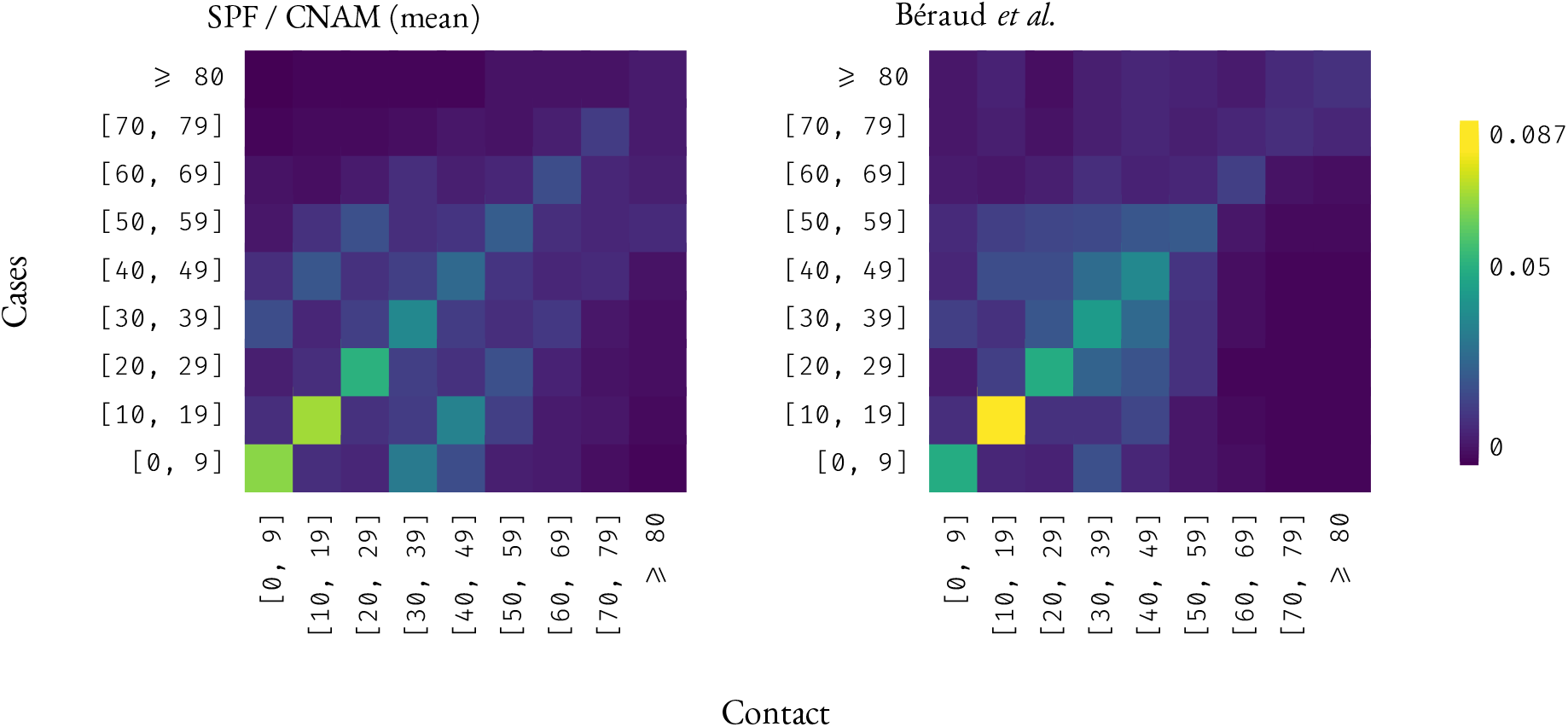
Contact matrices sources. The first source corresponds to 38 weekly contact matrices from August 2020 to April 2021 provided by SPF / CNAM (on the left, the mean contact matrix over the 38 weeks.). The second source originates from Béraud et al. (2015) (on the right). All matrices were normalized in order to be compared.

Therefore, we included all (normalized) contact matrices in the sensitivity analysis.

### Model outputs and fitting procedures

The main model outputs are population sizes of the compartments overtime for the year 2021 in France. The French publicly available hospital admission data are not stratified by age so we only use the global incidence data for parameterization and comparison purposes. In our model, incidence data in hospital admissions dynamics corresponds to the entry in the severe infection compartments (*I*^*s*^(*t, a, i*) and *I*^*d*^(*t, a, i*)) with a twelve days lag (Salje et al., 2020).

For each compartment dynamic, we build a 95% confidence interval using the 0.025 and 0.975 quantiles at each time step of all model runs used for the sensitivity analysis (see below).

Regarding parameter inference, we consider a daily minimal sum of squares between the data and simulations. We first fit the vaccination rate *ρ*(*t, a*) on French data as detailed in Supplementary Methods B.6. Due to high computational cost, we fit the NPI policies efficacy only on the median trajectory (defined as the trajectory obtained using the median parameter set but with the Béraud et al. (2015) contact matrix).

### Sensitivity analysis

We perform a variance-based sensitivity analysis to assess the robustness of the model given its inputs. We compute the Sobol main sensitivity indices for each model parameter and for each time step (Saltelli et al., 2008). For an input parameter *X*_*i*_ and a given day, this index reflects the fraction of the variance in the output *Y* (here the daily hospital admissions) and is defined by

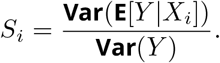

The difference between the sum of the main indices and 1 corresponds to the variance originating from the interactions between all the parameters. The analysis was performed on 30, 400 model runs with different parameters sets chosen using a Latin Hypercube Sampling within the ranges detailed in Table S1.

Assessing the sensitivity of model outputs depending on the contact matrix is more delicate since drawing each matrix coefficient would be numerically too costly and drawing an entire matrix would cause a loss of information regarding the role of the different age classes. However, we possess 38 weekly contact matrices from SPF and another contact matrix from Béraud et al. (2015). Therefore, for each age class, we randomly draw the corresponding age class column (*i*.*e*. the rate of being infected for the given age class by all the age classes) among the 39 available matrices. As discussed in Section, the two sources of contact matrices exhibit qualitatively different patterns, suggesting potential differences in terms of within-household transmission or active population transmission patterns. To avoid giving more weight to a specific pattern, the Béraud et al. (2015) matrix was weighted 38 times more than the SPF matrix.

Additional details regarding the sensitivity analysis can be found in Supplementary Results C.

## Results

### Basic reproduction number and NPIs

The basic reproduction number, denoted ℛ_0_, is a widely used metric in epidemiology because it corresponds to the average number of secondary infections caused by an infected host in an otherwise fully susceptible population (Anderson and May, 1992). Calculating it for our PDE system is not trivial and for this, we use the next generation operator approach (Diekmann et al., 1990; Inaba, 2012). More precisely, we show that the number of new infections in individuals of age *a* at time *t* in a fully susceptible population, denoted *I*_*N*_ (*t, a*), satisfies the renewal equation (see Supplementary Methods A.3 for details)

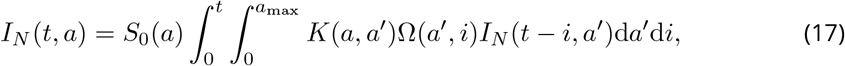

where Ω(*a, i*) can be interpreted as the infectiousness expectation of an individual of age *a* infected since time *i* and is defined by

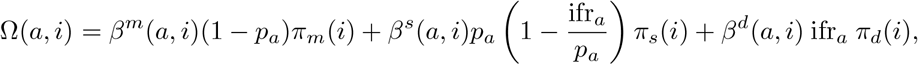

where *π*_*ℓ*_ is the “survival” probability (*i*.*e*. remaining in the compartment) of infected individuals of the *I*^*ℓ*^ compartment. Mathematically, 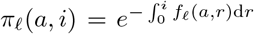, with *f*_*ℓ*_ = *γ*^*m*^, *γ*^*s*^, *μ*, respectively, for *ℓ* = *m, s, d*.

Following the Next Generation Theorem, the basic reproduction number ℛ_0_ is calculated as the spectral radius, noted *r* (*U)*, of the next generation operator *U* defined from 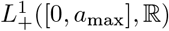, into itself by

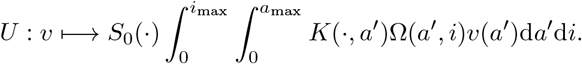

For parametrization purpose, we assume that the contact matrix *K*(·, ·) is given up to a positive constant *β* (to be determined), such that 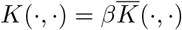, and 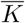 satisfies 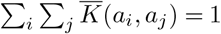. Consequently, we find that *β* is given by

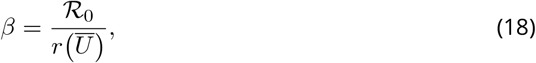

where 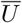 is the operator defined from 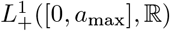 into itself by

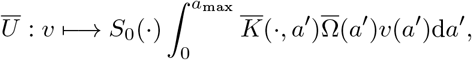

with

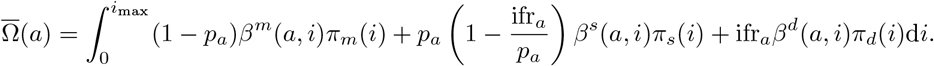

In the following, the ℛ_0_ is set to correspond to that of the *α* and then the *δ* VOC, which are both higher than that of the initial lineages. Note that, within this study, we scale *K*(·, ·) by

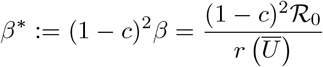

rather than *β* because we estimate the level of NPI efficacy (*c*) beforehand on real data, and for prospective scenarios after the current date, we arbitrarily set it to the desired value.

### Sensitivity analysis

Performing per time-point sensitivity analyses on the daily hospital admissions for all the model parameters (Figure 4), we noticed that most of the variance originated from the contact matrix (and its 9 parameters), especially the younger age classes. This effect was even more striking when considering the raw variance originating from each parameter (Figure S3).

**Figure 4.**
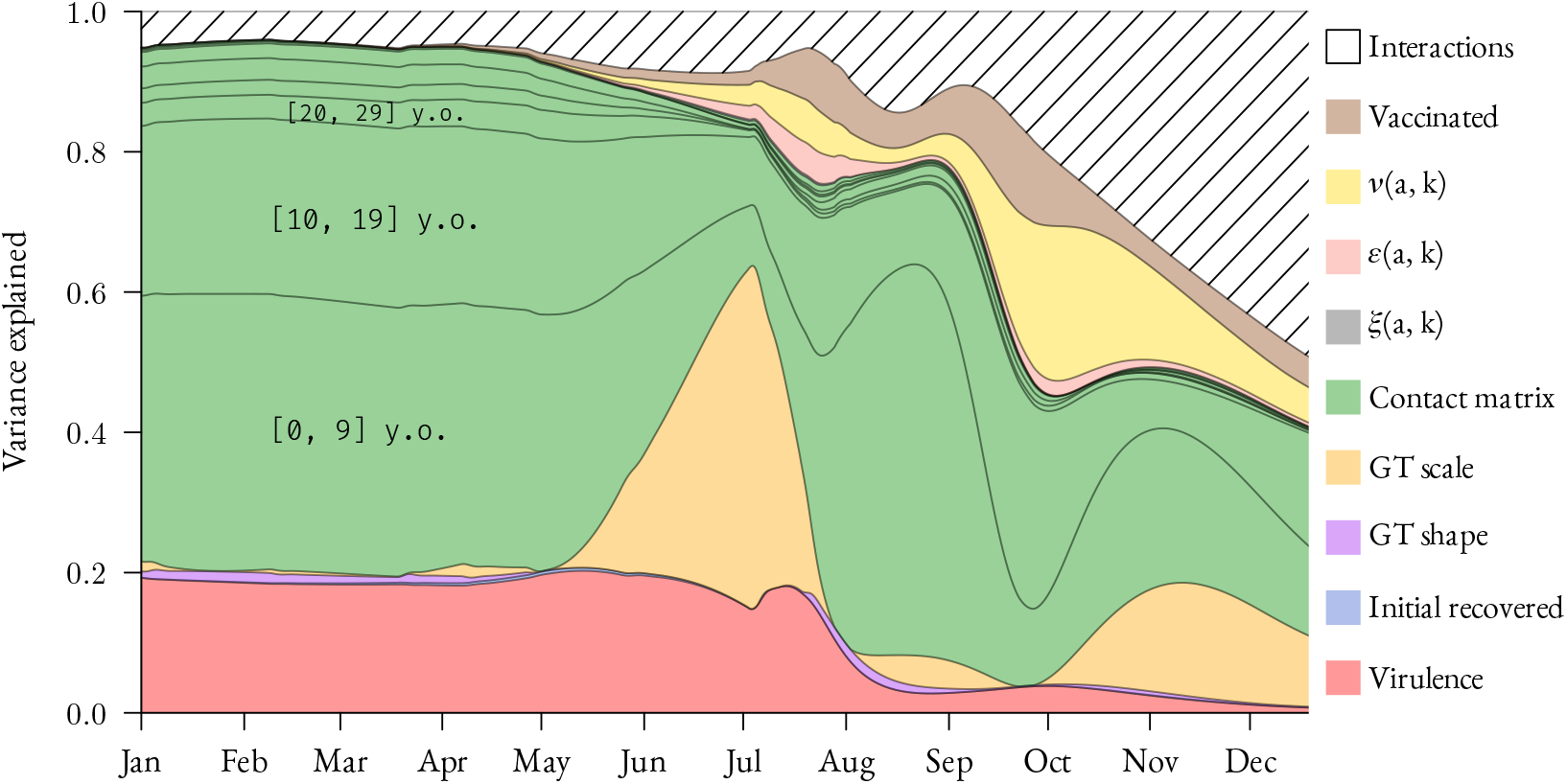
Temporal sensitivity analysis. We represent the main Sobol indices for each time step. These indices give the relative variance explained by each parameter. There are 9 parameters associated to the contact matrix corresponding to the rate of being infected for each age class from younger to older (bottom to top). ‘Virulence’ corresponds to the (increased) virulence of the VOC.

We also observed important time variations of some parameters, such as the generation time Weibull’ scale parameter. The period where this is the most predominant also corresponds to the period with few newly hospital admissions (Figure 6), and therefore a lesser variance. The sharp decrease of this parameter sensitivity coincides with the epidemic’s growth reprisal.

**Figure 5.**
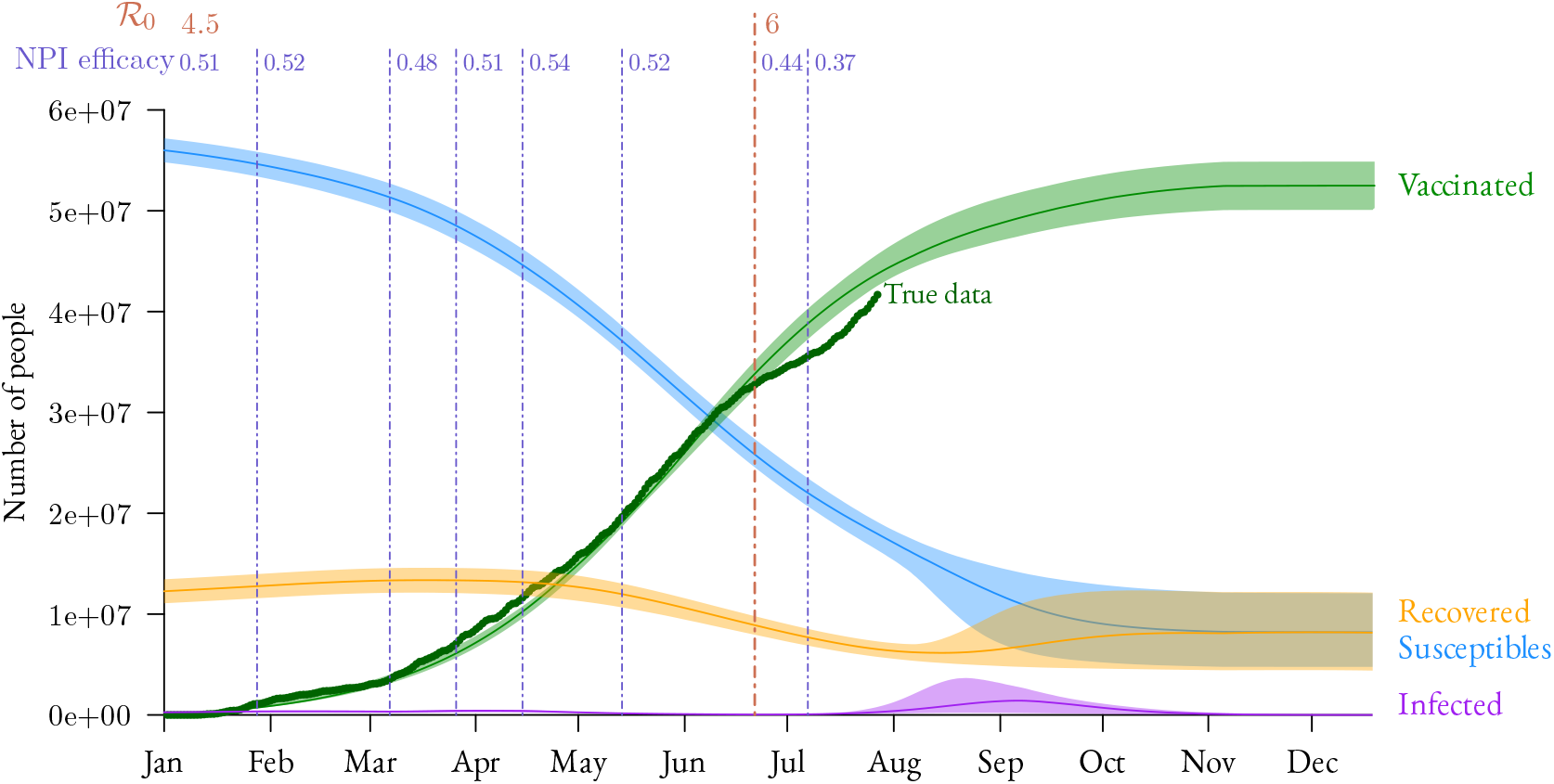
Epidemic dynamics of the French SARS-CoV-2 epidemic in 2021. For vaccinated hosts, we show the real data (green dots). Vertical lines indicate imposed changes in the basic reproduction number ℛ_0_ (in red) or estimated changes in the efficacy of NPIs (in blue). Plain lines and shaded areas respectively represent the median and 95% confidence interval computed from the simulations used for the sensitivity analysis.

**Figure 6.**
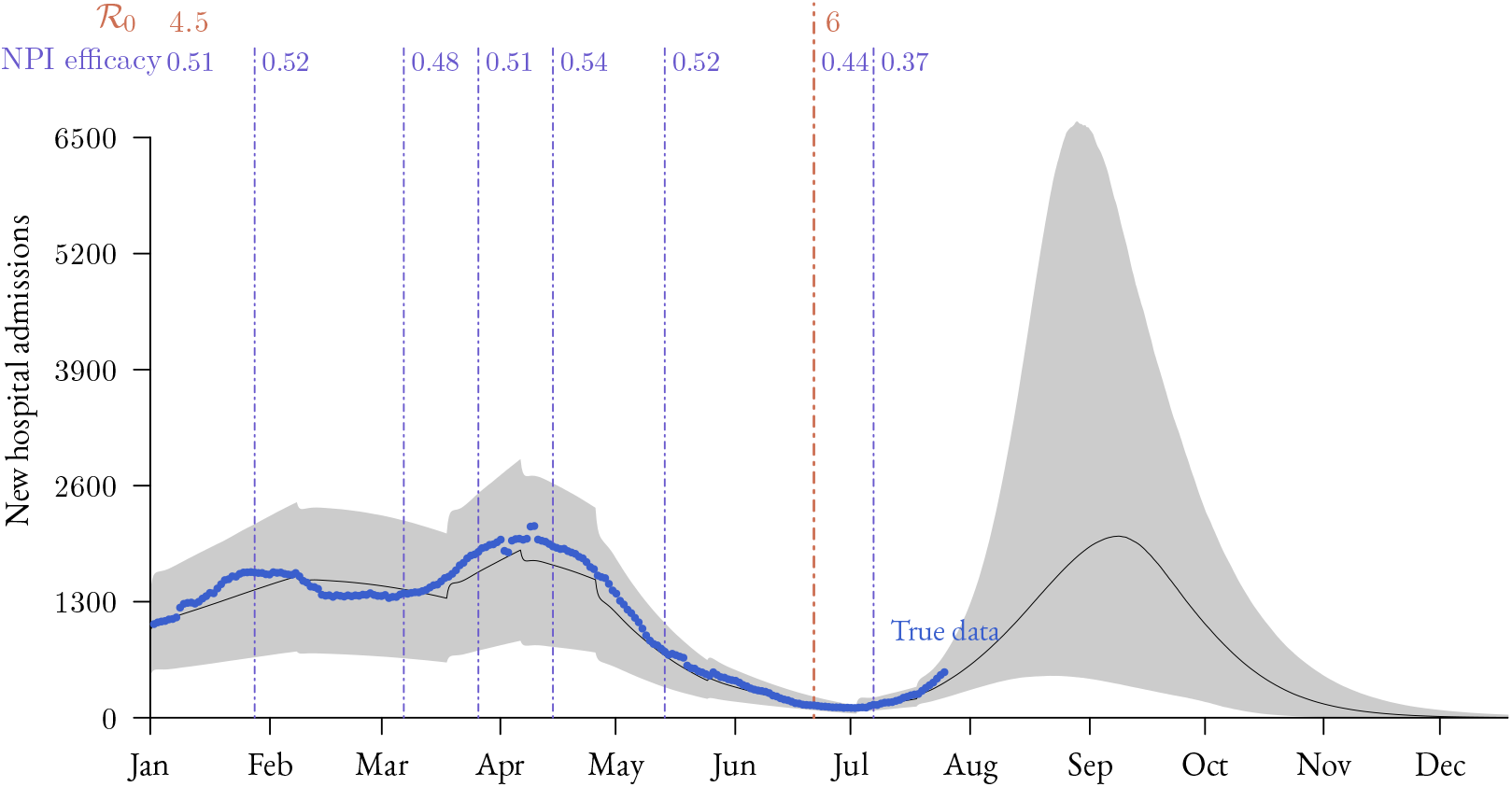
Daily admissions for Covid-19 in French hospitals in 2021. The blue dots show the real data, the plain line shows the median and the gray area shows the 95% confidence interval computed from the simulations used for the sensitivity analysis. As in Figure 5, vertical lines show the imposed and estimated values of ℛ _0_ and NPI efficacy (*c*).

Furthermore, others parameters explained variance, such as the VOC-increased virulence (in red) at first or more notably the interactions between parameters progressively increasing over time reaching half of the variance explained at the end of the year.

### Inferred dynamics

By parameterizing our model with existing data and inferring additional parameters, we could estimate past epidemic dynamics and investigate scenarios for future trends (Figure 5). The vaccinal coverage modelling did follow quite well real data, even though dissimilarities emerged in the summer (corresponding to the French summer holidays).

We may also observe a slight rebound in infected individuals mid-March, which follows by two weeks the end of the winter holidays. We also see this phenomenon on the new hospital admissions (Figure 6), with a supplementary delay corresponding to the delay between infection and hospital admissions.

It seems that the current vaccination rate is not high enough to avoid a new epidemic wave. However, we observe the uncertainty is huge and does not allow us to have a precise idea of what might happen.

## Discussion

Mathematical modelling has emerged as a central tool to control and anticipate the SARS-Cov-2 pandemic. This importance is likely to increase now that vaccination has become the cornerstone of the public health response in many countries. However, the limitations of current vaccination models lie in either neglecting memory effects or compensating by highly dimensional models with dozens of ordinary differential equations. In this study, we used partial differential equations to model medium and long term hospital admission dynamics in a population with natural and vaccine-induced immunity only with 8 general compartments.

To identify the components of our model that affected the results most, we conducted a global sensitivity analysis, which revealed that the contact matrix between age classes strikingly contributed more variance in daily hospital admissions than the VOC related increase of virulence itself. This predominant role is somehow surprising because although there is susceptibility to infection differences based on age (*e*.*g*. (Davies, Klepac, et al., 2020)), the strongest age differences appear in the IFR. Furthermore, in our results, contacts of younger age groups appeared to be the most important contributor to the variance of the outcome, although they were, and by far, the less likely to be hospitalized.

An important limitation of the model is that the contact matrix is assumed not to vary throughout a simulated epidemic. As suggested by the temporal variance in the SPF matrix data (Figure 2), this may be oversimplistic. For instance, we observed a difference of patterns in simulations whether they assumed high or low contact rates among younger age classes (as shown in Supplementary Figure S4). A baseline for the different contacts rates, if such a concept can even exist biologically, would most likely be impossible to determine because of the variety of events over a year inducing changes in social interactions such as calendar events (*e*.*g*. school holidays), implementation of control policies (*e*.*g*. lockdown, curfew), or even media coverage of the epidemic (resulting in spontaneous behavioural change with respect to the perception of the epidemic).

The importance of the age-structure of the host population in shaping Covid-19 epidemics is widely acknowledged. However, this effect is usually studied in the clinical context of disease severity and less so for transmission dynamics (Salje et al., 2020; Sofonea et al., 2021). However, there are exceptions and, using a PDE formalism, Richard et al. (2021) find the population structure to be the parameter that contributed relatively the most variance to their model’s output. Both Richard et al. (2021) and Keeling, Hill, et al. (2021) use a constant contact matrix, but they explore the impact of age-differentiated NPI policies. That being said, none of those studies (including ours), are able to fully assess the role of the age-structure since additional unknown patterns could potentially impact medium-term forecasting. For example, in absence of external data, it seems impossible to distinguish the “true contact matrix” from age-differentiated NPI policies. The problem is that the two would likely yield different outputs in NPI-lifting scenarios.

Although the variance contributed by the other parameters is low, there is a noticeable effect of the increase in virulence associated with infections caused by VOCs. Our approach allows us to quantify this effect (20 % of the variance) and even identify its peak contribution (in the declining phase of the epidemic peak). Note that the relative importance of virulence is lower in the prospective part of the model (*i*.*e*. after August 2021) but this is potentially because other parameters such as the ones related to vaccination and interactions between parameters become more important over time. We also did not consider an increase in virulence in the *δ* VOC compared to the *α* VOC (but recent data shows this might very well be the case (Sheikh et al., 2021)).

The generation time Weibull’s scale parameter, which has an impact on the mean generation time, also affects hospital admission dynamics, especially in June/July and November/December 2021. However, this needs to be put in perspective since the impact of this parameter arises only when there is little variance (Figure S3) and a decreasing epidemic (Figure 6). This can be explained by the fact a shorter mean generation time for a given reproduction number is known to increase the epidemic’s growth rate (Nishiura, 2010; Wallinga and Lipsitch, 2007). On this aspect, it is worth noting that the *δ* VOC seems to have a shorter generation time than the wildtype strain and this was not taken into account in this model (Zhang et al., 2021).

By applying our model to the context of the French epidemic, we show that the vaccination levels reached in the summer 2021 were insufficient to prevent a new epidemic wave, even in the scenarios with good vaccine coverage and efficacy (i.e. the lower bound of the confidence interval). A strong caveat to extending this model to longer time scales is that anticipating variations in the vaccination rate is extremely difficult as it relies on sociological factors. Increasing the uncertainty range for this parameter would most likely increase the variance in the sensitivity analysis. However, given past vaccination dynamics, we do not expect this to qualitatively affect the results. Regarding the medium-term forecasting, we did not include potential weather-related variations in behaviour or infectivity, which have been estimated to account for 15 to 20% of the variations in temporal reproduction number (Ma et al., 2021).

To analyse our model, we had to make several simplifying assumptions, which are common to differential equation-based models. The two major ones are the lack of spatial heterogeneity and the contact homogeneity among a given age class. The lack of spatial heterogeneity implies an identical contact rate across the whole country. This is not problematic at the start of an epidemic but is not adapted for long-term modelling as it affects the persistence of the disease (Hagenaars et al., 2004; Lloyd and May, 1996). Furthermore, age contact patterns allow us to capture some of the heterogeneity in the population but there could be other social heterogeneities that could, for instance, correlate with vaccination status. As shown in the case of the influenza virus, these could affect epidemiological dynamics (Barclay et al., 2014).

One advantage of this PDE model is the restrained number of compartments, especially compared to a classical alternative in ODE-based models which consists of chaining and multiplying compartments. For the latter, this would also require rewriting the formalism, depending on whether we consider a short, medium, or long-term temporal scale. In a way, PDE models allow one to explore a great variety of biological scenarios without adding any compartments thanks to the time since an event (infection, recovery, or vaccination) structure, only by varying the “age-since-event functions”. Therefore, the same model can be used to monitor new hospital admissions or the need for a new vaccination campaign years later in the presence of immune waning, *i*.*e*. a time-induced loss of immunity. For instance, we could account differential building up of vaccine immunity in susceptible versus recovered individuals. This would be consistent with the fact that the latter enter the vaccinated compartment at a later ‘vaccination age’ (*i*.*e. k* > 0) and a single vaccine dose appears to be sufficient to build strong immunity (Mazzoni et al., 2021). Also, in the context of waning immunity, our model can be used to investigate the long-term benefits or costs of implementing vaccine boosters depending on assumptions regarding vaccine efficacy or the duration of natural immunity. More generally, we can readily investigate the effect of implementing age-stratified vaccination policies.

Undeniably, PDE formalism requires a greater investment to implement simple models. Furthermore, deriving analytical results is more challenging, as illustrated by our calculation of the basic reproduction number. Another potential downside is that the computation time for simulation can increase rapidly.

On a more prospective side, our model offers promising possibilities to investigate virus evolution because it can explicitly capture the interplay between change in susceptibility, contagiousness, virulence and immune escapes (post-infection and vaccine) and trade-offs between them.

### Data, script and code availability

The model was implemented in `R` (v4.1.1) (R Core Team, 2021). The data and scripts used within this study are available in the repository https://gitlab.in2p3.fr/ete/covid19_vaccination_model as well as in the permanent Zenodo repository https://doi.org/10.5281/zenodo.5549752.

## Data Availability

Data and scripts are available in the linked git repository.

https://gitlab.in2p3.fr/ete/covid19_vaccination_model

## Acknowledgements

The authors acknowledge the ISO 9001 certified IRD i-Trop HPC (South Green Platform) at IRD Montpellier for providing HPC resources that have contributed to the research results reported within this paper.

We thank all the ETE modelling team for thoughtful discussions.

We thank the French public health agency (Santé Publique France) and the French national health insurance (Caisse Nationale d’Assurance Maladie) for providing Covid-19 contact matrices, and especially Alexandra Mailles and Jonathan Bastard.

BR is funded by the Ministère de l’Enseignement Supérieur et de la Recherche.

Version 3 of this preprint has been peer-reviewed and recommended by Peer Community In Mathematical and Computational Biology (https://doi.org/10.24072/pci.mcb.100008)

## Appendix

### Supplementary figures and table

**Figure S1.**
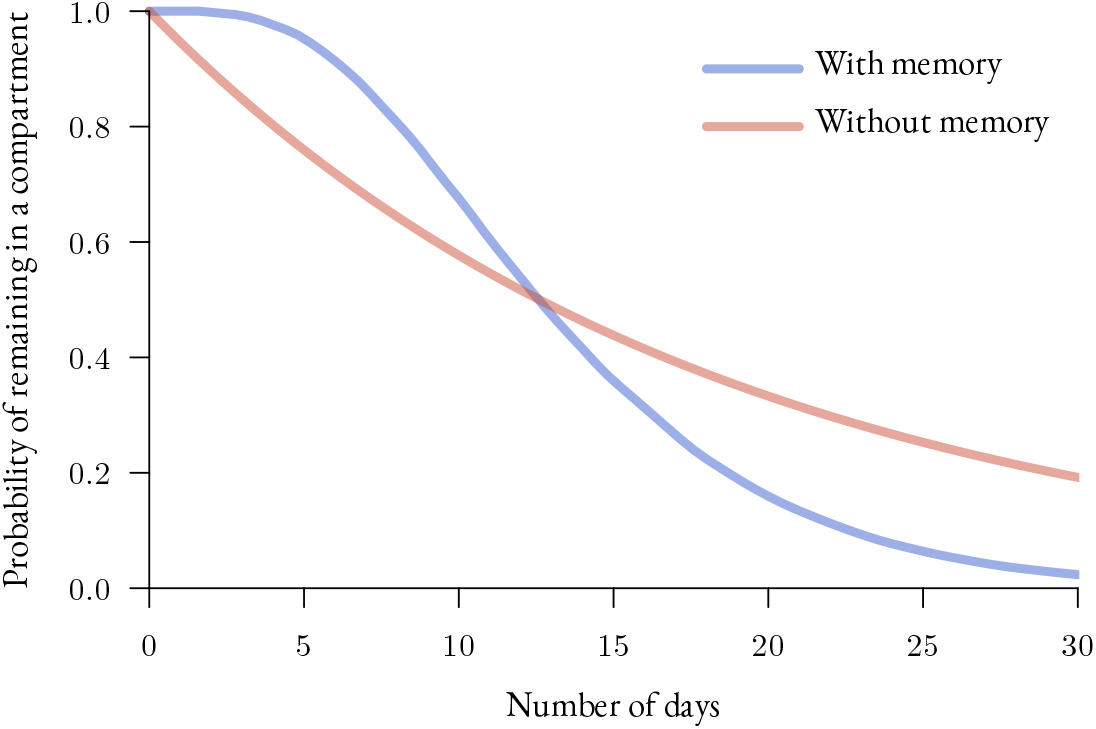
Illustrating the impact of memory on the time spent in a compartment. Providing memory to any compartment might yield different results in terms of epidemic dynamics. As illustrated here, in both cases half of the people leave the compartment at approximately twelve days. However, the number of people still in the compartment at 5 or 30 days might be radically different, given the initial population, whether memory is provided or not.

**Figure S2.**
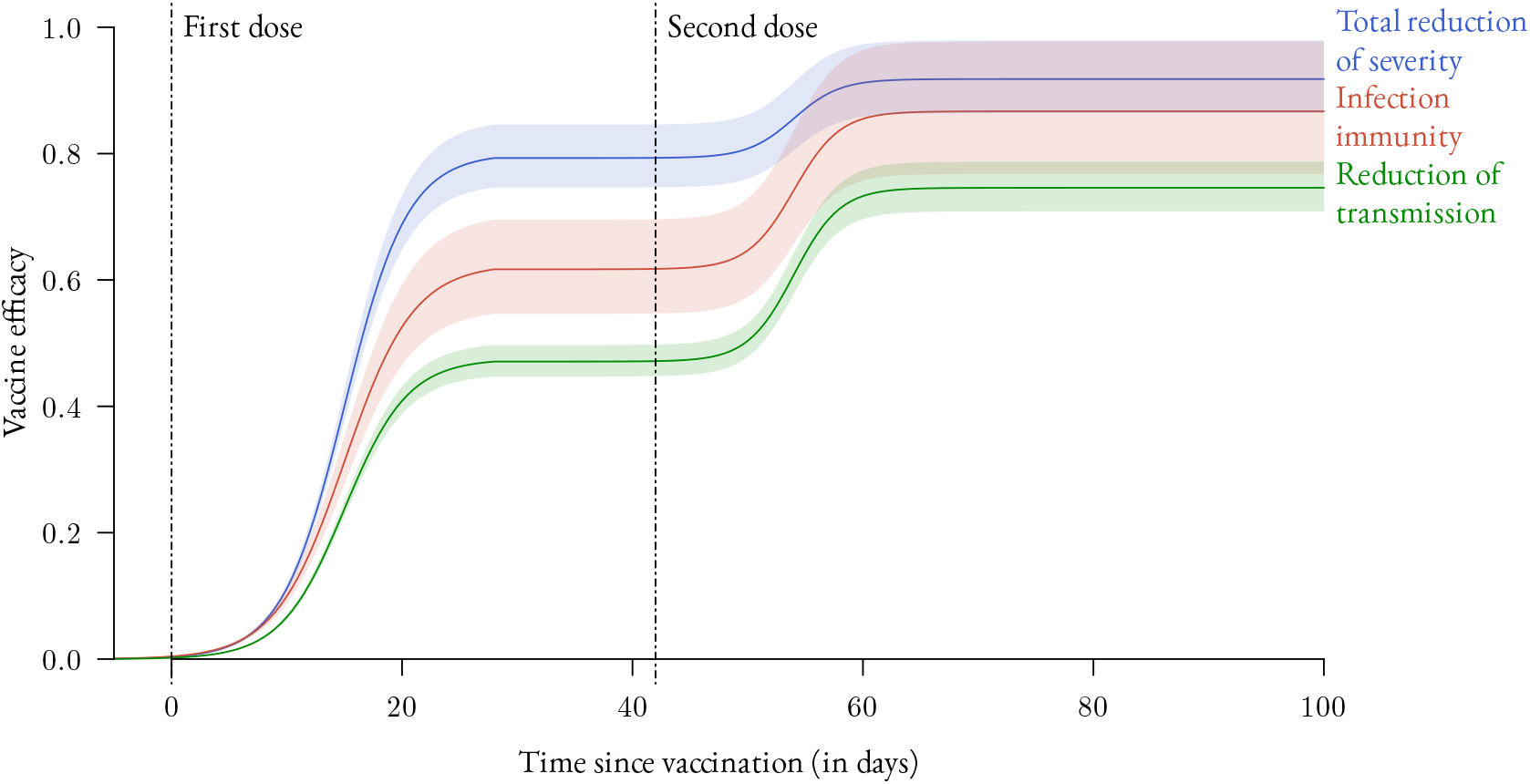
Temporal dynamics of vaccine efficacy. The double sigmoid is intended to reflect a two-doses vaccination schedule. The colors show the different types of protection conferred by the vaccine. The different efficacy levels remain constant over time after the second dose (no immune waning). Data used to calibrate these curves originate from the analyses of the Pfizer-BioNTech vaccine by Public Health England (2021). Note that the “Total reduction of severity” also accounts for infection immunity.

**Figure S3.**
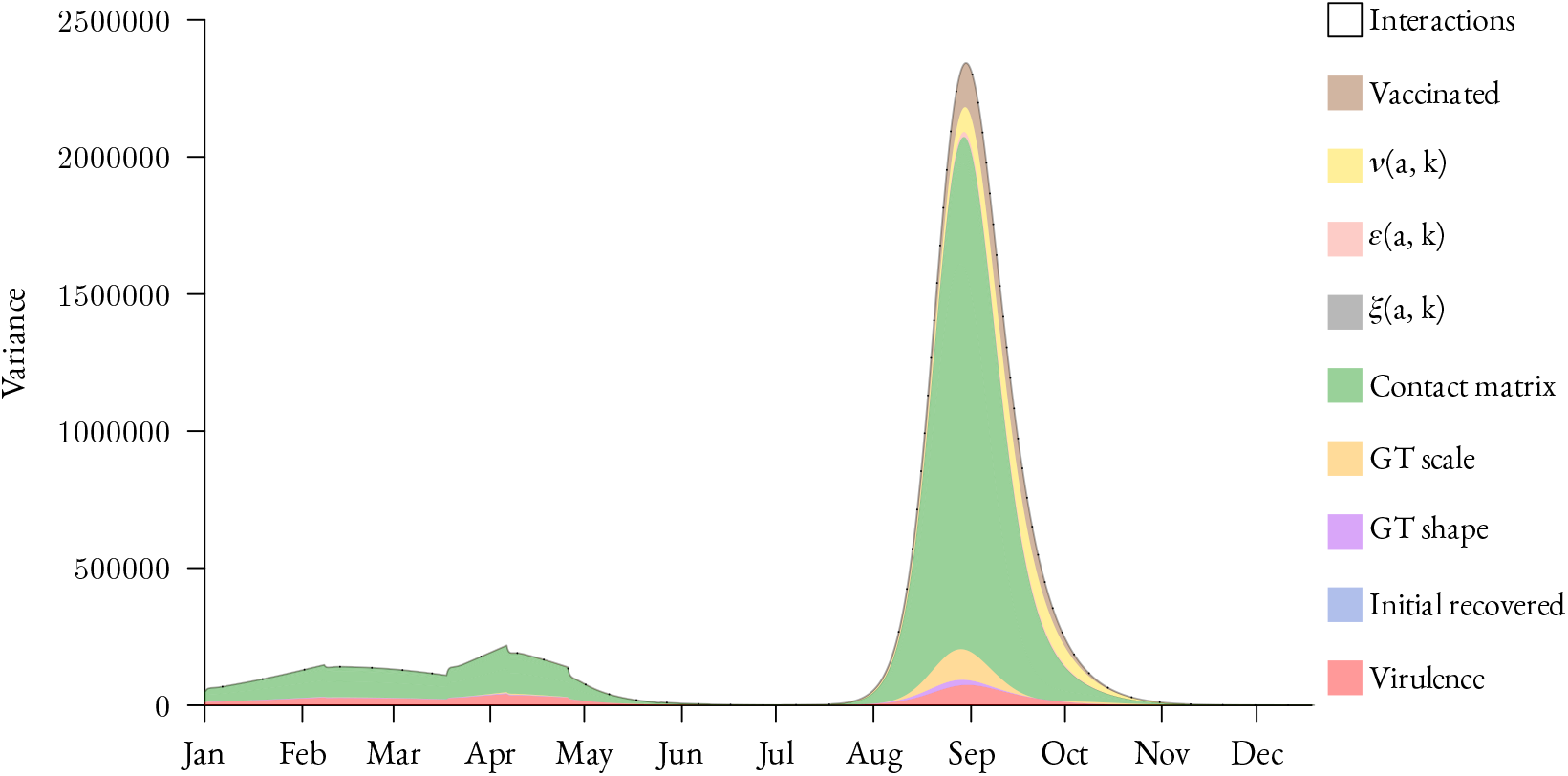
Non-normalised sensitivity analysis results. The output is similar to that in Figure 4 but we show, for each time step, the raw variance originating from the different parameters instead of the relative variance.

**Figure S4.**
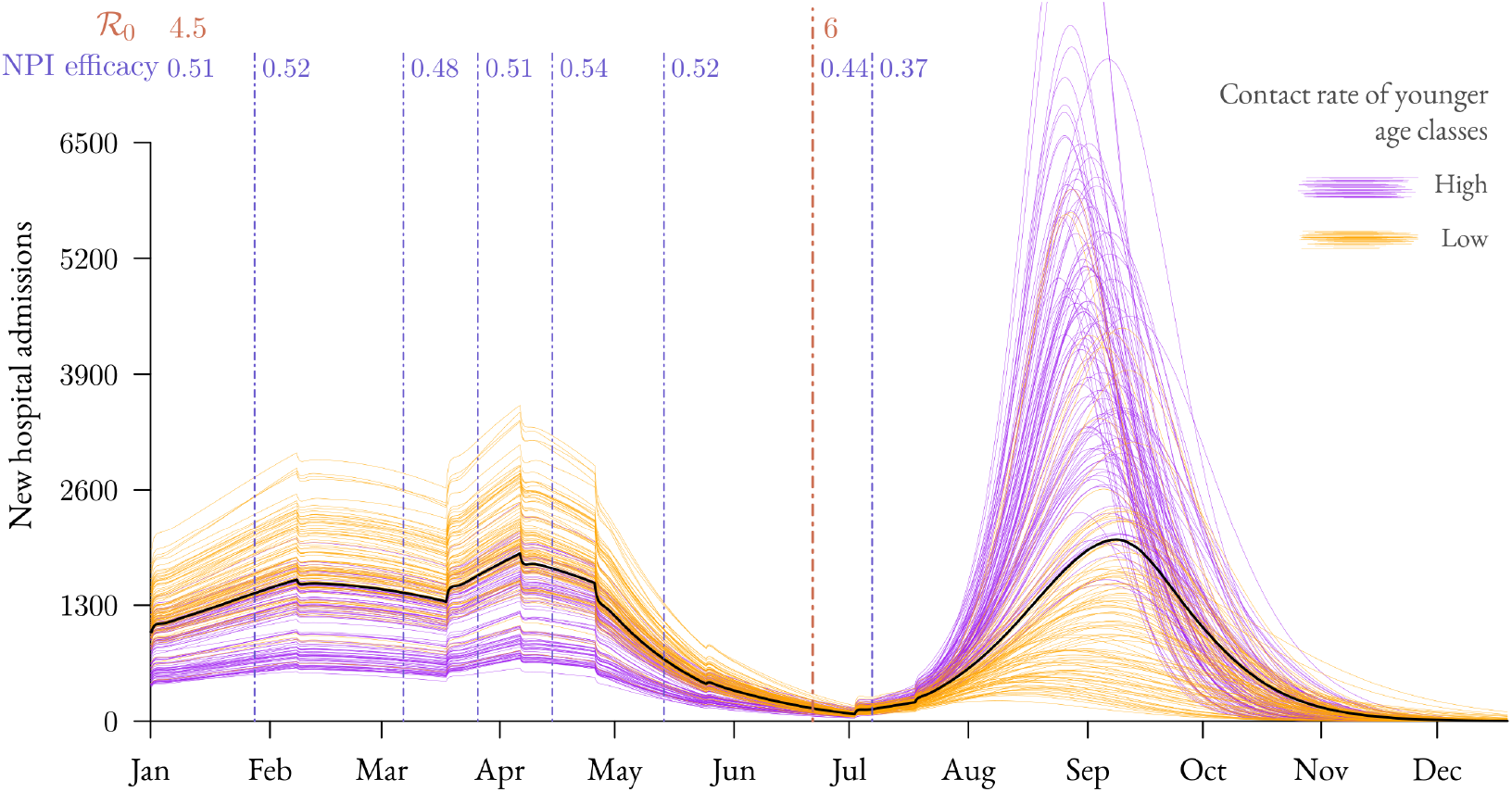
Younger age classes contact rates effect on hospital admission dynamics. We show 100 randomly selected trajectories among runs made with younger age-classes having the lowest (resp. highest) contact rates. We selected 50 trajectories among runs made with the 3 contacts matrices having the lowest (resp. highest) contact rates among [0 −9] y.o., plus 50 for the [10 − 19] y.o.

**Table S1.**
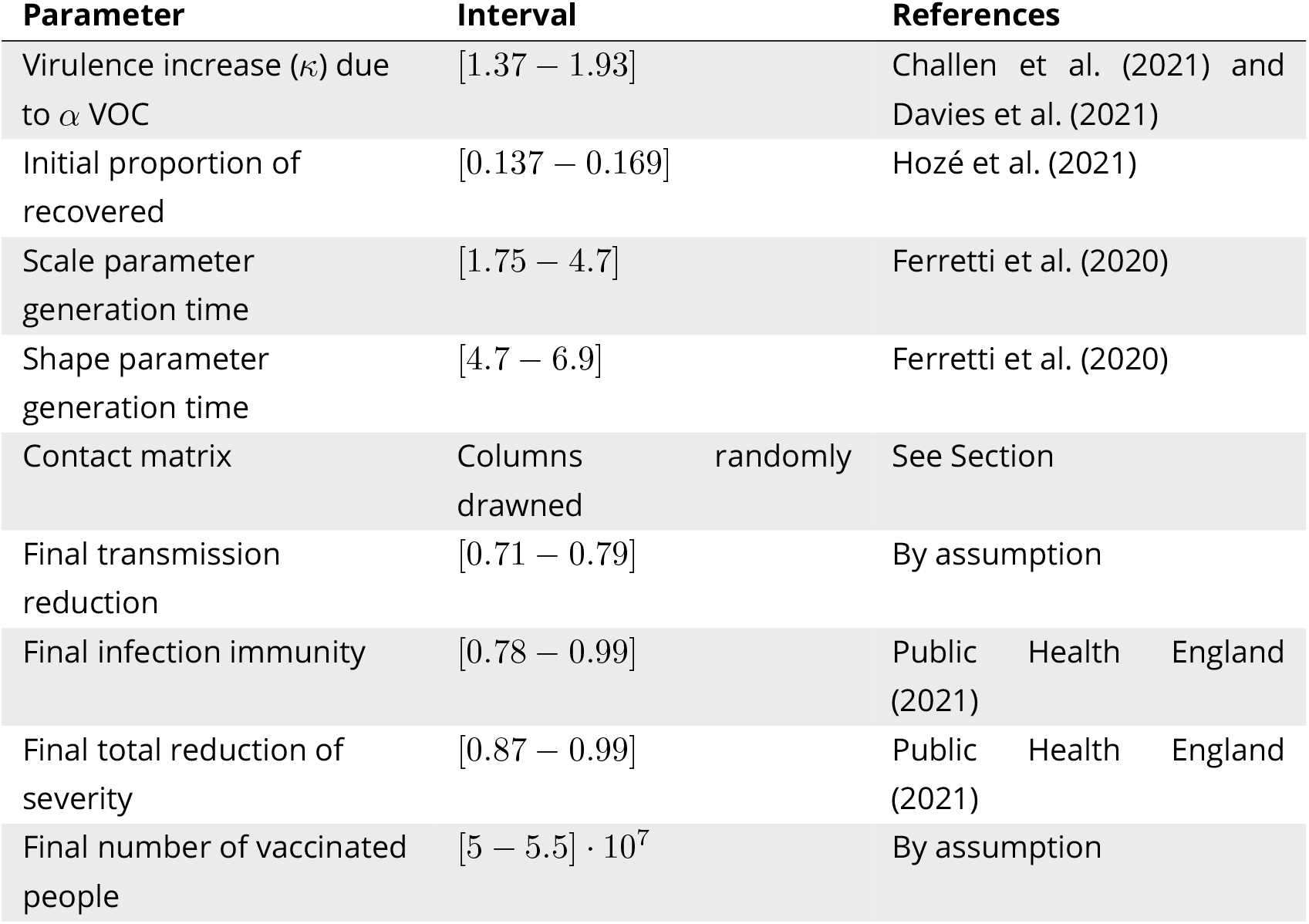
Parameters range used in the sensitivity analysis. Further explanations are provided in Supplementary Methods B, especially when assumptions are made.

### A Model

System (1)–(16) is considered under the following general assumption

#### Assumption S1

1. *p*_*a*_ ∈ [0, 1], *and* ifr_*a*_ ∈ [0, *p*_*a*_] *for all a* ∈ ℝ_+_;
2. 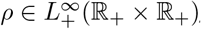, *with* 0 ≤ *ρ(t, a)* ≤ 1, *for all* (*t, a*) ∈ ℝ_+_ × ℝ_+_;
3. 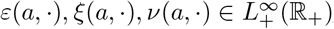, *for all a* ∈ ℝ_+_, *and* 0 ≤ *ε*(*a*, ·), *ξ*(*a*, ·), *ν*(*a*, ·) ≤ 1;
4. 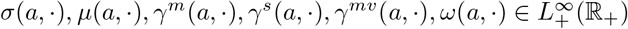, *for all a* ∈ ℝ_+_;
5. 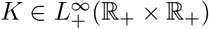;
6. *Transmission rates satisfy* 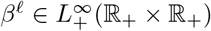 *for each ℓ* ∈ {*m, s, mv*}.

#### A.1 Implementation

The model was implemented in `R` (R Core Team, 2021), using Rcpp (Eddelbuettel and François, 2011) to maximize computational efficiency.

The PDE system was implemented using an Euler explicit scheme.

#### A.2 Well-posedness

Let us introduce the Banach space as well as its positive cone

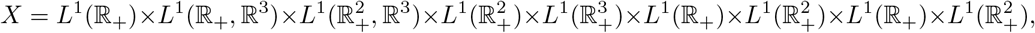

as well as its positive cone

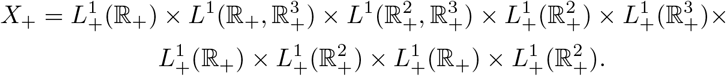

Now, we define the subspaces 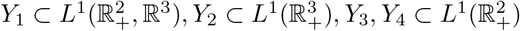 by:

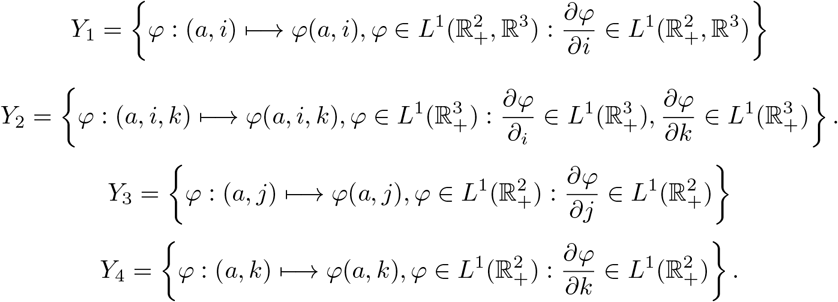

It follows that there exists a unique linear operator 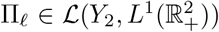 for each *ℓ* ∈ {1, 2} such that Π_1_*ψ* = *ψ*(·, 0, ·) and Π_2_*ψ* = *ψ*(·, *·*·, 0) for all *ψ* ∈ *Y*_2_. Next, let *A* : *D*(*A*) ⊂ *X* → *X* be the linear operator on the domain

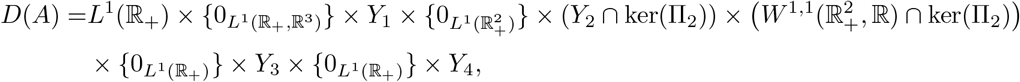

and defined by

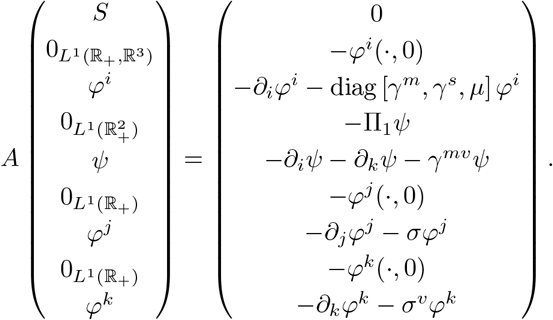

Finally, let us introduce the following nonlinear map 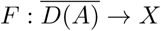:

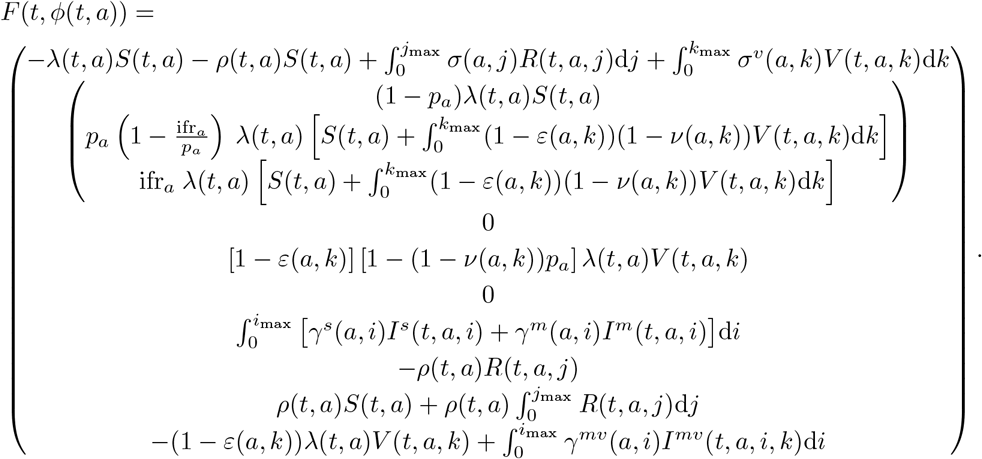

wherein *ϕ*(*t*) is the function:

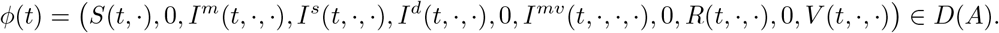

From here, System (1)–(16) rewrites as the following nondensely defined Cauchy problem:

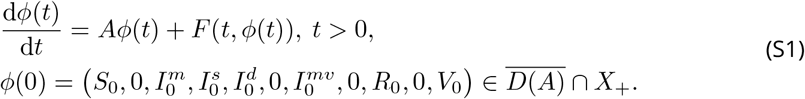

Therefore, under Assumption S1, we have the well-posedness of System (S1); that is, the Cauchy problem (S1) generates a unique globally defined, positive and bounded non-autonomous semiflow.

The proof of this result is based on a rather standard methodology combining an intgrated semigroup approach and Volterra integral formulation in the context of multiple structured variables (*e*.*g*., Richard, Choisy, et al. (2022) and the references therein) and existence of the semiflow for non-autonomous systems (*e*.*g*., (Magal, 2001; Pazy, 2012)).

#### A.3 Basic reproduction number and NPI policices

We consider that there are no vaccinated, *i*.*e ρ*(*t, a*) ≡ 0 and *V* (*t, a, k*) ≡ 0, nor recovered people, *i*.*e. R*(*t, a, j*) ≡ 0, and that the number of susceptible individuals is very close to the total population size. For simplicity, we first introduce the “survival” probability (*i*.*e*. the probability to remain in the compartment) of infected individuals of, respectively, the *I*^*m*^, *I*^*s*^, and *I*^*d*^ compartments:

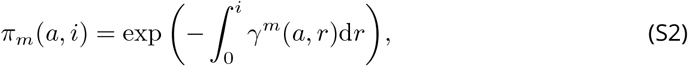

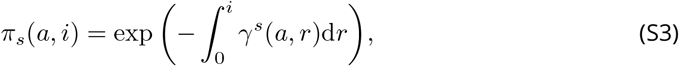

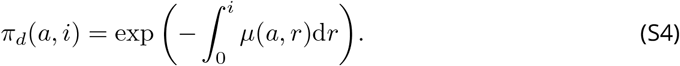

By linearizing System (1)–(16), we obtain the following Volterra formulation for *I*^*m*^, *I*^*s*^, and *I*^*d*^ compartments:

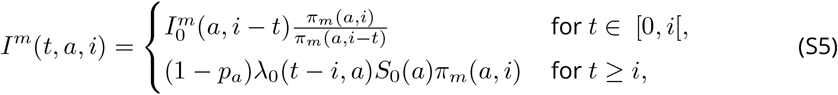

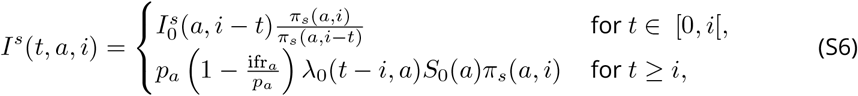

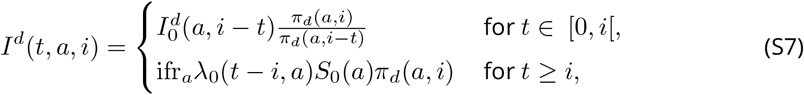

with *λ*_0_(*t, a*) defined as *λ*(*t, a*) with no control policies (*c* = 0),

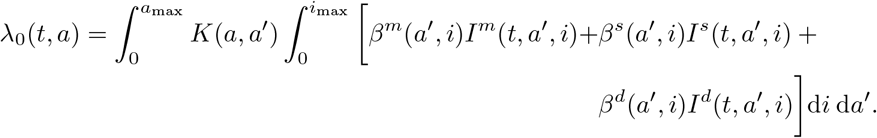

Let *I*_*N*_ (*t, a*) = *λ*_0_(*t, a*)*S*(0, *a*) be the density of newly infected of age *a* at time *t*. Then, by (S5)–(S7) it comes

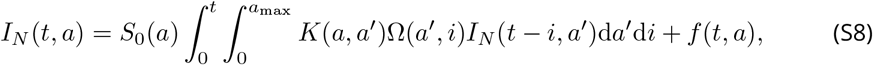

where

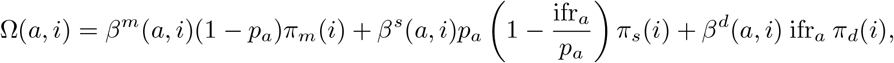

and *f* (*t, a*) is accounting for the initial population.

The basic reproduction number ℛ_0_ is then the spectral radius, denoted by *r*(*U*), of the next generation operator *U* defined from 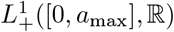 into itself by

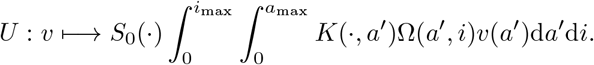

For parametrization purpose, we assume that the contact matrix *K*(·, ·) is given up to a positive constant *β* (to be determined), such that 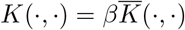, and 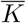 satisfies 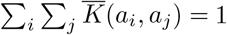. Consequently, we find that *β* is given by

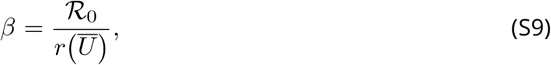

where 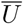 is the operator defined from 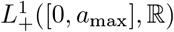 into itself by

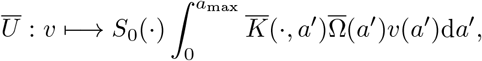

with

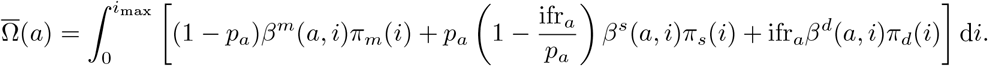

Note that, within this paper, we scale *K*(·, ·) by

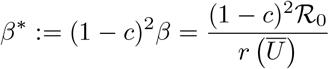

rather than *β* since the NPI level efficacy was fitted beforehand on real data.

To go further steps in the computation of 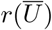, in addition to the general Assumption S1, we also assume that

##### Assumption S2

*Functions S*_0_, *K*, 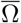 *are positive almost everywhere*.

Then, we can show that 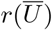 is given by the spectral radius of the following linear operator, defined from 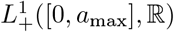 into itself:

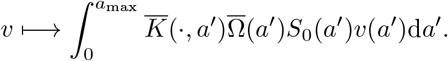

The spectral radius of this later operator is computed easily since the age *a* is numerically divided into *n* ∈ ℕ^*^ classes so that the term inside the integral of the latter equation is a *n* × *n* matrix. Finally, the scaling parameter *β* is obtain from (S9).

Importantly, the symmetric property of the contact matrix *K* is not strictly necessary for the computation of 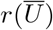. However, in addition to Assumptions S1 and S2, if *K* is a symmetric function, then the Rayleigh quotient formulation leads to (see Proposition F.2 in Richard, Alizon, et al., 2021)

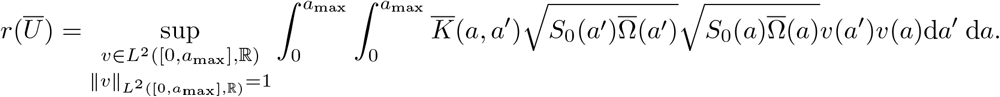

### B Model parametrization

In this section, we describe the parametrization and the assumptions made in the main text. The uncertainty ranges retained for each parameter are displayed in Table S1.

#### B.1 Proportion of severe cases, IFR and increase in virulence

The proportion of severe cases corresponds here to the fraction of the population who will be hospitalized following a SARS-CoV-2 infection. This parameter is age-dependent and follows the infection fatality rate (IFR) by Verity et al. (2020).

However, studies show that the virulence of the infection increased (taken into account by the *κ* parameter) by more than 60% with the *α* VOC (Challen et al., 2021; Davies et al., 2021). Estimation for the *δ* VOC are still at an early stage, so we used the virulence from the *α* VOC, even if the former seems more virulent (Sheikh et al., 2021).

#### B.2 Generation time and transmission rates

For the underlying generation time, we use that provided by Ferretti et al. (2020), which follows a Weibull distribution, with a shape parameter of 2.826 (95% CI [1.75 − 4.7]) and scale parameter of 5.665 (95% CI [4.7 − 6.9]). We assume that the transmission rates from mild infections *β*^*m*^(*a, i*) and severe cases *β*^*s*+*d*^(*a, i*) are equal to the generation time corrected by the probability for individuals to leave their infected compartment.

#### B.3 Recovery rates

From the data shown in Salje et al. (2020), we retrieve the time severely-infected individuals spend as infected, whether they required ICU admission or not, by adding up the different exponential distributions of the different infected compartments of their model (which will be denoted *E*_1_, *E*_2_, *I*^hosp^, *I*^non ICU^, *I*^ICU^, *H*_1_, *H*_2_, *H*_ICU_, *ICU* _1_, *ICU* _2_ in a later study by Kiem et al. (2021)). This gives us the probability of remaining in the infected compartment over time, thereby allowing us to infer the recovery rates. We also know from Lefrancq et al. (2021) the probability for hospitalized individuals to require ICU admission, which provides us with appropriate recovery rates weighting for severe cases.

We apply the same method for mildly-infected individuals.

#### B.4 Initial conditions

For this PDE model, the initialization is not as straightforward as for ODE models since within a compartment individuals do not have the same age of infection (for infected individuals) or time since clearance (for recovered individuals). Initialising over all the domain of definition of each compartment is difficult since a uniform initialization would almost immediately be counterbalanced by a higher probability to leave the compartment for higher ages (*i*.*e. i* > 0 and *j* > 0). Put differently, this would produce distributions different from what we might expect with a constant inflow in the compartments. To overcome this issue, we start the different runs with a 45 days delay (not shown) to let the different compartments stabilise around a distribution.

Hozé et al. (2021) estimate that the proportion of recovered adults in France was of 0.149 (95% CI [0.132 −0.169]) on January 15th, 2021. For simplicity, and in absence of more detailed data, we assumed this proportion to be constant across age classes, including the younger age groups.

For infected individuals, we initialize the density with a qualitative value. The *β* coefficient associated to the ℛ_0_ and the NPI were fitted such that the number of daily hospital admissions was close to the real data. The overall number of infected individuals in the model on January 1st was around 267, 000 [170, 000 − 310, 000].

#### B.5 Vaccine properties

We assume that the three types of vaccine efficacies (against infection, severe forms, and transmission) follow a double-sigmoid temporal pattern starting from the day of injections (Figure S2). The reduction in transmission rate corresponds to function *ξ*(*a, k*) in our model, the infection immunity corresponds to function *ε*(*a, k*). The total reduction of virulence corresponds to the cumulative effects of *ε*(*a, k*) and *ν*(*a, k*).

The order of magnitude of the final (full) efficacy levels is based on that from the Pfizer-BioNTech vaccine after two doses provided by Public Health England (2021). However, note that the outcomes referenced in the report were used as overall proxy in this study (“symptomatic disease” and “infection” for *ε*(*a, k*), “hospitalisation” and “mortality” for *ν*(*a, k*) and “transmission” for *ξ*(*a, k*)) as they do not exactly match our implementation. The reduction of transmission estimation 14 days after the second dose was not available, so we applied a rule of thumb in order the increase between the two injections was similar to the other vaccine properties. Hence, we assume a 75% efficacy for transmission reduction 14 days after the second dose.

We also assume there was no difference in efficacy between age classes, and that the different efficacy levels remain constant 14 days after the second dose until the end of our projections (*i*.*e*. no immune waning).

#### B.6 Vaccination rate

We model the vaccination rate using a sigmoid function,

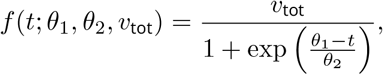

where *v*_tot_ denotes the total number of vaccinated individuals at the end of the year (which is a model input), and *θ*_1_ and *θ*_2_ are the sigmoid curve parameters fitted to the observed data. This gives us the number of newly vaccinated people at each time step.

The number of doses attributed to each age-group at each time step depends on initial weights (*ω*_*a*_(*t*_0_)), which can be interpreted as the age-based strategy vaccination prioritization, the proportion of the age group targeted (*t*_*a*_), assuming that the total number of vaccinated in each age class may vary and is lower than 100%, and the proportion of individuals already vaccinated within each age-group at time *t* (*p*_*v*_(*t, a*)).

Therefore, at time *t* +Δ*t*, the splitting of the number of doses is given by *ω*_*a*_(*t* +Δ*t*) which is defined by

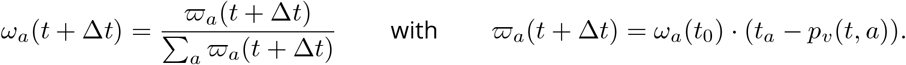

Finally, we have

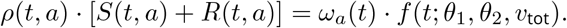

The initial weights, *ω*_*a*_(*t*_0_), and *t*_*a*_ is fitted on an ordinary least squares metric to reproduce at best the real vaccination rate.

Vaccination is assumed to start on January, 1st, 2021.

#### B.7 Age groups

The different data sources we used had non-homogeneous age groups, and these groups sometimes overlapped. For instance, the contact matrices were provided by 5 years bins, while the parameters related to the disease severity were provided by 10 years bins. On another hand, vaccination data were provided with age groups better reflecting the French society structure (0-4, 5-11, 12-18, 18-24…).

We decided to use 10 years bins age groups since it was the option that required less data transformation.

### C Sensitivity analysis

To perform the sensitivity analysis, we use the lhs package (Carnell, 2020) to generate a Latin Hypercube Sample (LHS). The parameters were drawn in a uniform distribution within the confidence interval specified for each parameter and shown in Table S1.

For each parameter combination, a model run was computed. In total, 30, 400 model runs were performed. Then, for each time step, we used the multisensi package (Bidot et al., 2018) to compute the Sobol main indices, given by

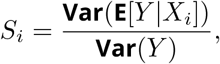

as implemented in the sensitivity package (Iooss et al., 2021). The difference between the sum of all the main indices and 1 corresponds to the effect of interactions between parameters. More explanations are available in Saltelli et al. (2008).

Due to numerical approximations, some indices may sometimes be negative (the lowest was −0.004). These were rounded to 0.

